# Machine Learning and Micro Capture-C resolve GWAS associations revealing endothelial stress pathways in Coronary Artery Disease

**DOI:** 10.64898/2025.12.18.25342557

**Authors:** Matthew Baxter, Edward Sanders, Simone G Riva, Joseph C Hamley, E Ravza Gur, James L T Dalgleish, Isabella M Freund, Nigel Roberts, Gabrielle Raymond, Martin Sergeant, Damien J Downes, Hangpeng Li, David G McVey, Shu Ye, Christopher Grace, Lance D Hentges, Gwendal Dujardin, Tom R Webb, James O J Davies, Theodosios Kyriakou, Anuj Goel, Hugh Watkins, Jim R Hughes

**Author notes:** These authors contributed equally (Edward Sanders, Simone G Riva).

## Abstract

Resolving the gene targets of non-coding genetic variation is the major bottleneck in translating genome wide association studies into mechanistic understanding of complex diseases such as coronary artery disease (CAD). Combining new Transformer-based Machine Learning (ML) approaches trained on cardiovascular epigenetics with high-resolution, allele-specific genomic and transcriptomic technologies we create a highly scalable platform to simultaneously resolve causal variants, cell-type of action, output gene, and direction of effect. When applied to CAD genetics, our ML predicts causal variants from 20,747 candidate SNPs across 9 vessel cell-types and identifies disrupted transcription factor binding motifs using ML feature attributions. We investigate 94 of the top predictions in endothelial cells using Micro Capture-C, revealing the importance of fluid shear stress and TGF-β signaling pathways. We exploit allelic skew in heterozygous cells to demonstrate both variant causality and effect direction, demonstrating this platform can be used to rapidly resolve non-coding genetics in complex disease.

## Introduction

Cardiovascular disease remains the primary cause of death worldwide ^1^. 50% of coronary artery disease (CAD) risk is genetic, and GWAS studies have identified hundreds of genetic loci which are significantly associated with disease risk ^2^. However, despite nearly two decades since the first GWAS studies for CAD, few of these statistical associations are translating into a better understanding of the affected genes and pathways and so to drug targets or therapies ^3^. This is undoubtedly due to the complexity of the systems that they alter. However, recent advancements in single cell genomics, Machine Learning (ML), and chromosome conformation capture (3C) technologies offer an exceptional opportunity to finally resolve these genetic associations at scale and to more quickly identify new therapeutic targets ^4–6^. There are four main challenges to solving a genetic association: i) identifying the effector cell type; the majority of linked variants are in the non-coding genome where they are considered to alter cell-type specific gene regulation ^7–9^. ii) identifying the causal variant; this is challenging because any given genetic signal normally encompasses 10s-100s of variants in high linkage disequilibrium, any of which may be the causal variant and where identification is also dependent on having accurately identified the correct cell type in which to determine this ^10,11^. iii) identifying the output gene; understanding which gene or genes are affected is highly challenging ^12^. iv) determining the direction of the effect as variants can both increase and decrease the functional output of the gene.

All of these challenges arise because GWAS variants overwhelming lie within the highly dynamic non-coding genome where the majority of gene regulatory elements function differentially across cell types and states ^13,14^. The major genomic regulatory elements that encode this cell and state specificity are enhancer elements ^15^. Enhancers may be located over a million base pairs away from their target genes and can lie much closer to unrelated genes or even buried in their introns ^16^. Fundamentally, enhancer elements are composed of combinations of specific transcription factor (TF) binding sites, which allows them to function in a cell-type specific manner, based on the specific combinations of TFs a given cell type or cell state expresses ^17^. Small changes to the genetic sequence of an enhancer can dramatically change the binding efficiency of a given TF or even create brand-new binding sites. Therefore, genetic variants located in enhancers have the potential to alter gene regulation through altered TF binding kinetics and so contribute to inherited genetic traits including disease susceptibility ^18^.

To tackle the challenge of identifying causal non-coding variants in a cell-type specific manner we have developed a Transformer-based Deep Neural Network trained on open chromatin data that can predict which variants within the genetic haplotype will alter the activity of specific regulatory elements within these cell types ^19^. We use this Machine Learning tool to prioritise candidate causal variants in coronary artery cell types for subsequent mechanistic investigation and gene network discovery.

Determining which genes are altered by such regulatory genetics has been a major block in the interpretation and exploitation of common genetics. However, our current understanding that enhancers and promoters come into proximity during gene activation led to the rapid development of multiple classes of chromatin conformation capture (3C) assays that used this property to try and map regulatory interaction in the genome ^6,20^.

The gene(s) which enhancers are regulating in a given cell type can now be resolved at base-pair resolution, and at scale, using Micro-Capture-C (MCC) ^21^. MCC is currently unparalleled in its ability to resolve enhancer-promoter interactions and importantly can be multiplexed to simultaneously resolve hundreds to potentially thousands of interactions within genetic loci ^6^. Two further properties of MCC make it the most effective approach for resolving the effects of enhancer variants: i.) protein binding footprints can be determined to delineate the TFs controlling gene expression and how this is altered in the presence of a variant; and ii.) heterozygous enhancer alleles can be directly compared within the same cell to accurately measure the effect of a variant on regulatory interactions, cell-type specifically and within the same sample to avoid confounding technical variation. This allele specific analysis, both at the level of TF footprints and at the enhancer-promoter interactions allows the direct and sensitive determination of causality for a given genetic variant within the same assay that also determines the likely affected target gene(s). MCC therefore provides us with a powerful and scalable solution to the third challenge of identifying the target genes of genetic regulatory variants while simultaneously providing further support for the cell type specific causality of the variants and implied directionality of the effect. To address the last challenge and confirm both the effect on the transcriptional output of these MCC linked genes and the directionality of the change, we use phased genome builds from long read sequencing combined with intron retaining RNA-seq (POINTseq) ^22^. This approach takes advantage of the naturally occurring variants in the introns of genes to measure the effect on nascent transcription of the allele interacting with the causal enhancer variant relative to the unaffected allele. This provides a highly scalable approach to address the final challenge, both confirming the target gene causality and the direction of the effect in primary cells without need for low throughput, challenging and often confounding experiments such as modelling variants using CRISPR editing.

We therefore present a novel experimental pipeline to resolve GWAS associations into the cell types, genes and pathways underlying these genetic traits. This pipeline identifies cell types enriched for the genetics under scrutiny, identifies causal SNPs in distal cis regulatory elements, the output genes through which they exert their effects and the directionality of these effects. We apply this pipeline to CAD genetics in endothelial cells, identifying affected genes at scale and highlighting shear stress pathways and TGF-β signalling as key effectors of CAD genetics.

## Results

### Machine Learning predicts causal SNPs in enhancers

To address the challenge of identifying causal SNPs in complex loci, we employed ML to prioritise candidate causal variants by evaluating the potential of each variant to disrupt cis-regulatory elements. Publicly available scATAC-seq data for diseased and non-diseased coronary arteries was used for transfer learning to fine-tune the REnformer ML platform ^4,19^. The cells were clustered into endothelial, smooth muscle cells (SMC1 & SMC2), pericytes, fibroblasts, macrophages, mast cells, B cells and T cells (Figure 1A). Pseudo-bulk ATAC tracks reconstructed from the reads within these clusters were used to train REnformer to predict cis-regulatory elements in each of these cell types. REnformer was able to predict cell type specific ATAC tracks with high accuracy (Figure 1A & Supplementary Figure S1). Because the predictions are based solely on DNA sequence, we were able to predict the effect of sequence variation on cis-regulatory elements in each of the coronary artery cell types (Figure 1B). To curate a list of possible causal SNPs, we performed FGWAS using coronary artery EC & SMC using summary data from Aragam et al 2022 and included variants with PPA>50% supplemented with genome-wide significant lead SNPs & 1% FDR variants (Supplementary Table 1). Each of these candidate causal SNPs was tested in the REnformer model against all of the coronary artery cell types to identify cell-type specific cis-regulatory elements that could be functionally affected by these sequence variations (Figure 1B). Both loss of function and gain of function SNPs were predicted in all cell types which can be browsed in the supplementary table (Supplementary Table 2).

**Figure 1.**
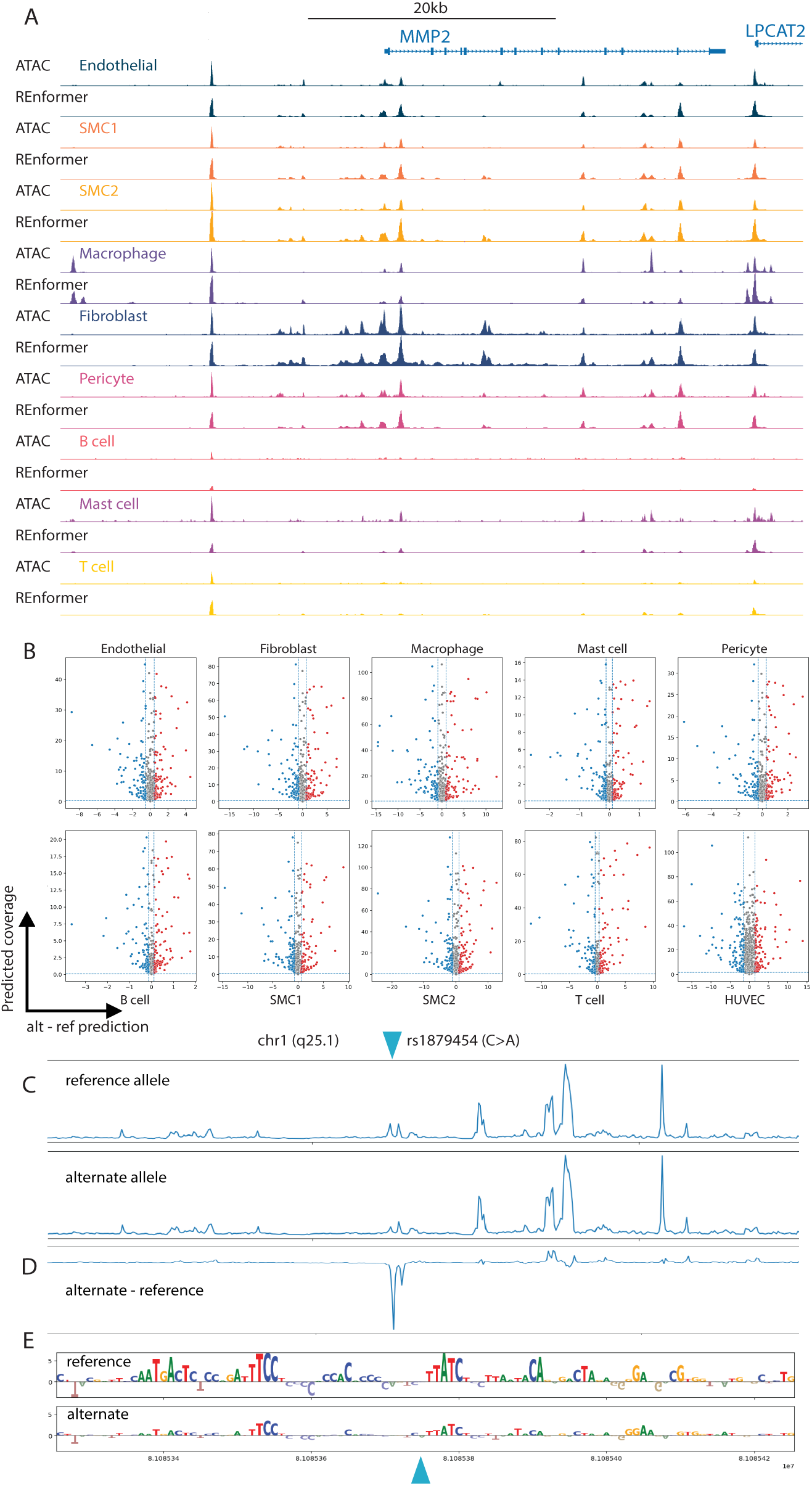
Machine Learning predicts the effect of genetic variants in coronary artery cell types. A. REnformer was trained on publicly available scATAC data from primary coronary artery tissue samples. REnformer was then used to predict the genome accessibility for 9 coronary artery cell types. An example locus around the MMP2 gene is shown. The tracks show the ground truth scATAC data (labelled “ATAC” followed by the cell type), and directly below the REnformer prediction for that cell type. B. The effect on genome accessibility of 21,348 CAD associated lead SNPs and high linkage disequilibrium candidate causal SNPs is predicted by REnformer across 9 coronary artery cell types as well as HUVECs. Predicted coverage (Y-axis) shows the predicted chromatin accessibility at a given SNP position. Alt-Ref prediction (X-axis) shows the predicted effect size of the candidate CAD SNP. Empirical significance cut-offs of 0.01 were calculated for each cell type; variants below the cut-off (loss of function) are coloured blue, variants above the cut-off (gain of function) are coloured red. C. Example REnformer predictions at the chr1:q25.1 locus for SNP rs1879454 (reference allele = C, alternate allele =A). The position of rs1879454 is shown by a blue triangle. D. The prediction difference for rs1879454 where the reference allele has been subtracted from the alternate allele. E. Feature attribution for the REnformer predictions in panel C. The relative contribution of each base pair in the reference and alternate genome to the prediction is shown by height (positive or negative) of the relevant letter. The variant position is shown by a blue triangle.

An example REnformer prediction is shown in Figure 1C,D. We extracted the prediction for CAD SNP rs1879454 (C/A) which was recently reported to regulate *TLNRD1* by disrupting a GATA/TAL binding site in an enhancer, and is the only functionally solved CAD endothelial enhancer variant to date ^23^. REnformer predicted the endothelial cell chromatin accessibility around the SNP rs1879454 for both the reference (C) and alternative (A) allele (Figure 1C), predicting a strong decrease in the enhancer accessibility with the A allele (Figure 1D), recapitulating the published observation. Furthermore, we examined the feature attribution from the REnformer prediction which showed a strong down-regulation of the GATA/TAL motif (CTTATCT), together with reduced motif activity across the enhancer, as a result of the single base pair substitution (Figure 1E). These observations show that REnformer is able to accurately identify causal enhancer SNPs in a cell-type specific manner, and identify the underlying transcription factor binding motifs which are affected.

### Prioritisation of candidate causal SNPs for mechanistic investigation

Because enhancer-promoter interactions cannot be predicted from chromatin states alone, we employed Micro-Capture-C to identify which gene(s) the prioritised enhancers interact with. Endothelial cells were chosen due to their previously reported involvement in the pathogenesis of CAD and the availability of HUVECs as a well-characterised and tractable cell model. ATACseq was performed on 3 independent HUVEC cell lines to ensure a range of genotypes, before merging the data from all 3 to identify regulatory elements. These elements were combined with public data for HUVEC histone modification (H3K4me1, H3K4me3, and H3K27ac) and CTCF occupancy to classify regulatory elements as enhancers, promoters, CTCF sites or intermediate states (Supplementary Figure S2).

REnformer predictions were then generated for candidate CAD SNPs lying within endothelial enhancers. For example, rs604723 is located within an intronic enhancer in the *ARHGAP42* gene body. REnformer predicted a strong loss of enhancer strength with the alternate C allele compared to reference T allele at position chr11:100739815 (Figure 2A). Interrogation of the ML model indicates that the T allele creates a “TTAC” motif which negatively affects the enhancer prediction. Conversely, for rs2508619 which is found in an intergenic enhancer at position chr11:75419204, REnformer predicts a strong gain of function with the alternate C allele compared to the reference T allele. In this case, examining the model reveals the creation of a “TTCCT” binding site for ETS transcription factors such as ETV6 (Figure 2B). Interestingly, a number of the commonly affected motifs in this analysis involved transcription factors reportedly involved in TGF-β signaling, for example rs582384 affects a SMAD/NFKB motif and rs17293632 affects a FOS/JUN motif (Figures 2 C&D). In total we prioritised 94 SNPs for further investigation with MCC. Capture probes were designed for these SNPs. The full prioritisation and probe design can be viewed here: https://mlv.molbiol.ox.ac.uk/projects/multi_locus_view/8712.

**Figure 2.**
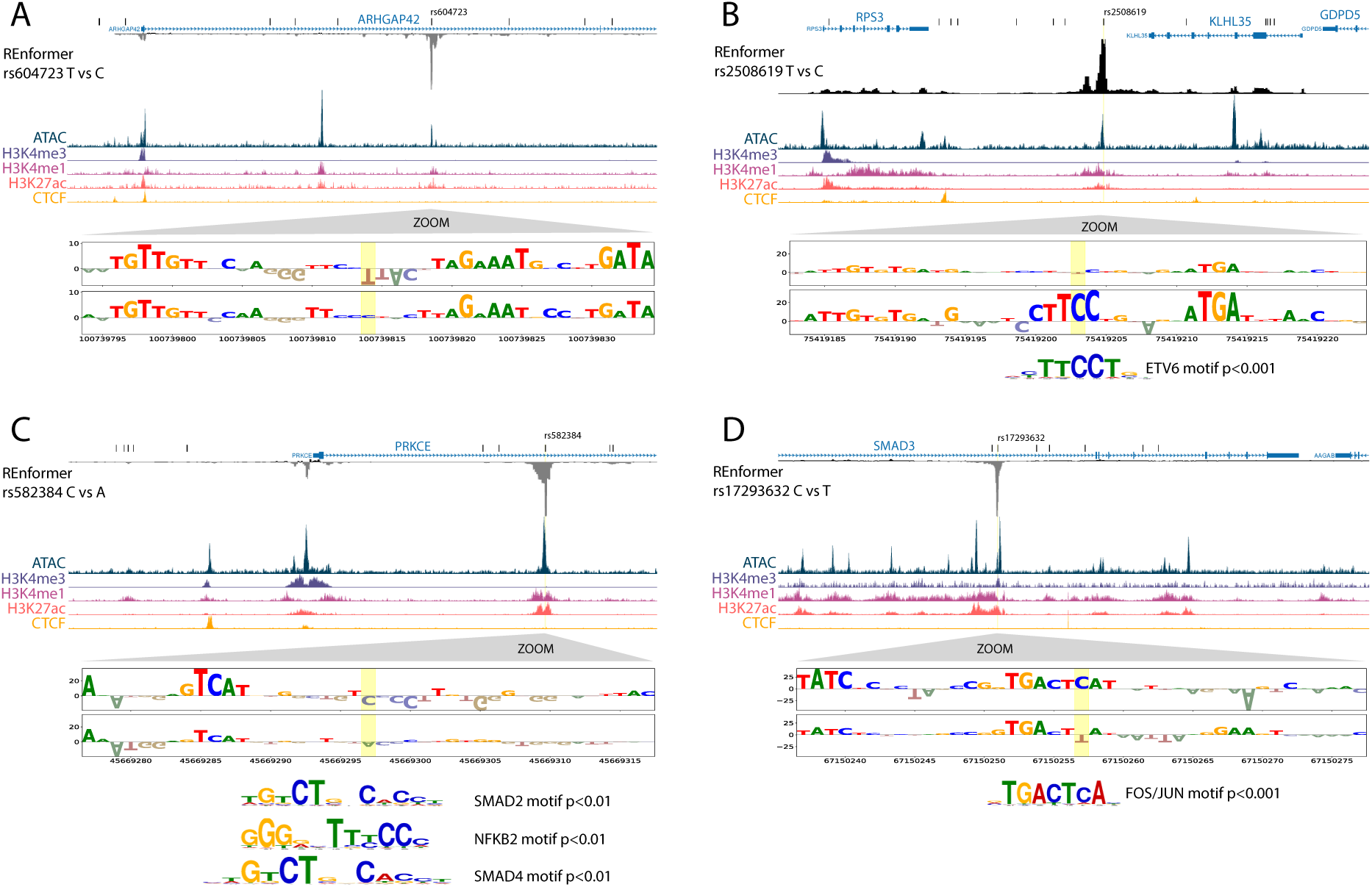
REnformer prioritises candidate causal variants in HUVECs. REnformer predictions are shown for 4 example candidate causal variants: rs604723 (A), rs2508619 (B), rs582384 (C), rs17293632 (D). In each panel, the prioritised SNP is labelled above the gene annotation, and other non-prioritised SNPs are shown as vertical black lines. The REnformer prediction (alt-ref) is shown below the gene annotation and directly above ATACseq data for HUVECs. Publicly available ChIPseq tracks (H3K4me3, H3K4me1, H3K27ac, CTCF) are also shown to aid identification of the genome regulatory elements. REnformer feature attribution for the 40bp around the prioritised SNP (yellow highlight) is shown below the genome tracks in each panel; the reference allele feature attribution is shown above the alternate allele. Statistically significant transcription factor binding motifs are shown below the feature attribution in B, C, and D. The identity of the transcription factor binding the affected motif in A is unknown.

### Micro Capture-C links enhancer variants to output genes

To link the candidate causal SNPs to output genes we generated MCC libraries for 3 independent HUVEC donors. Libraries were captured with probes specific to the 94 prioritised enhancers and interaction networks visualised in UCSC. An example locus is shown in Figure 3A. This is a highly complex locus containing 71 candidate causal SNPs in high linkage disequilibrium with two independently associated CAD lead variants (rs11466359 and rs60315715). REnformer predicted strong loss of function for two SNPs 24 base pairs apart in the same enhancer (rs2241718 and rs2241719), which are in linkage disequilibrium (R>0.8) with both lead variants. Importantly, this prediction was consistent across both coronary artery endothelial cells and HUVECs (Supplementary Table S2). We designed a capture site spanning these two candidate causal SNPs which are located in an enhancer downstream of the *TGFB1* gene, and is annotated in Gencode as the 3’UTR of *CCDC97* (Figure 3, blue triangle).

**Figure 3.**
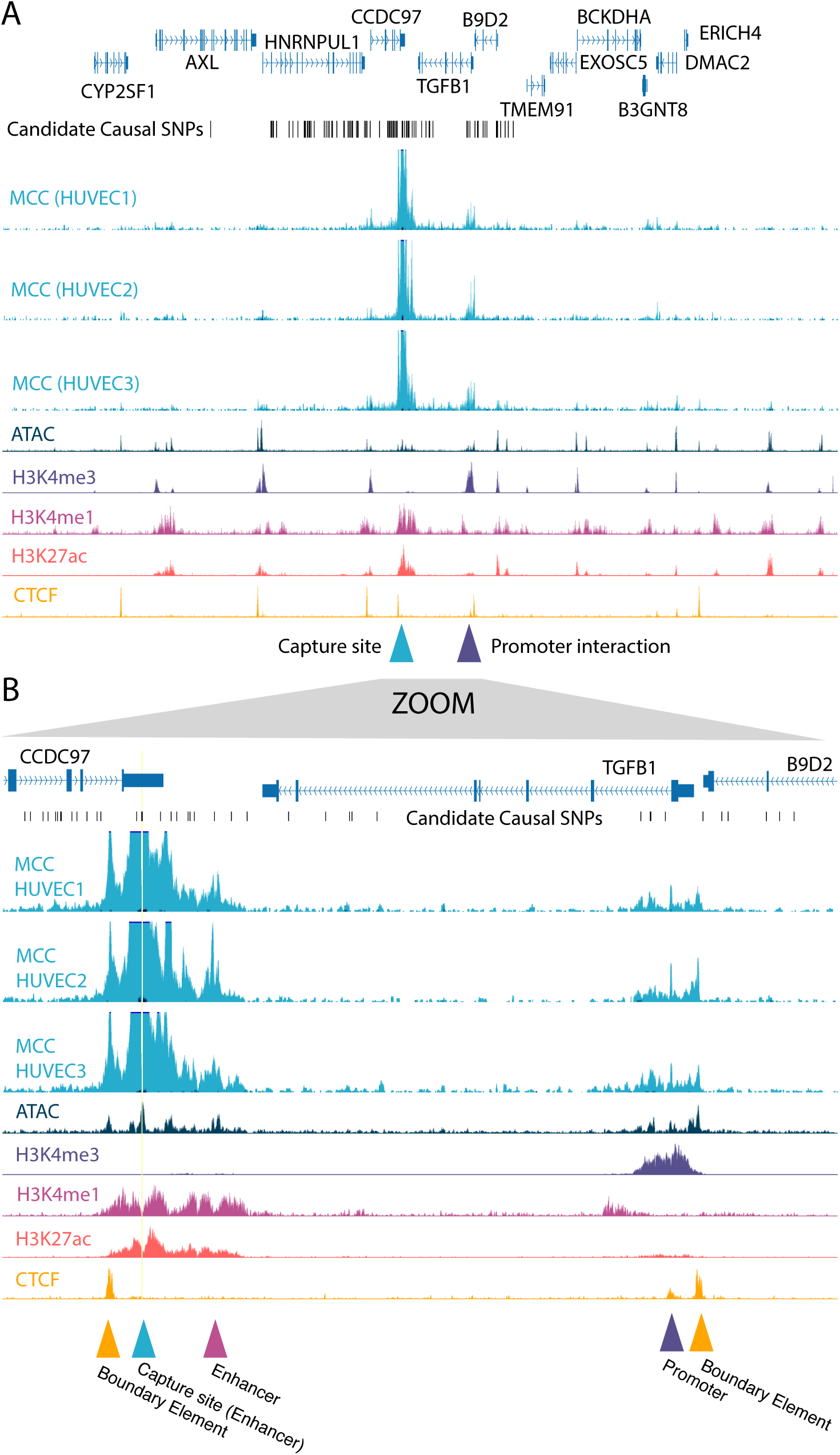
Micro Capture-C links candidate causal variants to output genes. MCC was performed on 94 enhancer elements containing candidate causal variants. Panel A shows the wider genomic context including lead and candidate causal SNPs and 12 genes in the locus. Three MCC tracks from 3 genetically independent donors are shown with the capture site located in the enhancer which intersects with candidate causal SNP rs2241718. A promoter interaction from this enhancer is indicated by the purple triangle below the tracks. Panel B shows a zoomed in view of the enhancer promoter interaction. CTCF boundary elements are indicated by yellow triangles, the capture site is indicated by the blue triangle, enhancer-enhancer interactions are indicated by the purple triangle, and the interaction of the captured enhancer with the promoter of TGFB1 is indicated by the purple triangle.

The top 3 tracks in Figure 3A show the MCC interactions for each of the 3 independent HUVEC donors. A strong peak in all 3 of the MCC tracks can be observed at this capture site, reflecting successful capture of the targeted enhancer. Approximately 20kb to the right, a smaller peak is observed which is aligned with the promoter of *TGFB1*. This indicates that the captured enhancer interacts, in this cell type, with the promoter of *TGFB1*. Importantly, no other strong promoter interactions are observed, indicating selectivity of this enhancer for the *TGFB1* promoter over the other 11 genes within the locus. Figure 3B provides a zoomed in view of the capture site and the promoter interactions, where more detail can be observed. A strong enhancer-enhancer interaction (pink triangle) is observed as well as the enhancer-promoter (purple triangle) interaction with *TGFB1*. The interactions are bounded by CTCF sites (yellow triangles), which may explain the specificity of this enhancer for *TGFB1*. The other 94 MCC capture loci can be viewed here: https://genome-euro.ucsc.edu/s/mdehxmb7/CAD_HUVEC_extended_data

### MCC interaction map highlights key endothelial functions in the pathogenesis of CAD

In total, 98 genes were found to be interacting with captured enhancers across the 3 HUVEC donors. These genes are visualised in the STRING network (Figure 4A) and were enriched for gene ontological terms including response to fluid shear stress, TGF-β receptor signalling pathway, regulation of cell migration, cellular response to stress, and regulation of cell migration (Figure 4B & Supplementary Table 5). Genes known to be important in the response to shear stress included the transcription factors *KLF2* and *KLF4*, as well as the mediators *TGFB*1 and *MMP2*. Key nodes central to cellular stress responses also emerge, coalescing around the cell cycle regulator *CDKN1A*. Other genes are important in regulating cell migration, such as *PRKCE* and *PLPP3*, which may reflect the importance of the role of endothelial cell signaling in smooth muscle cell and fibroblast migration to the progression of atherosclerosis. The MCC links between candidate causal SNPs, lead SNPs, and interacting genes are summarised by HUVEC donor in Supplementary Table 3.

**Figure 4.**
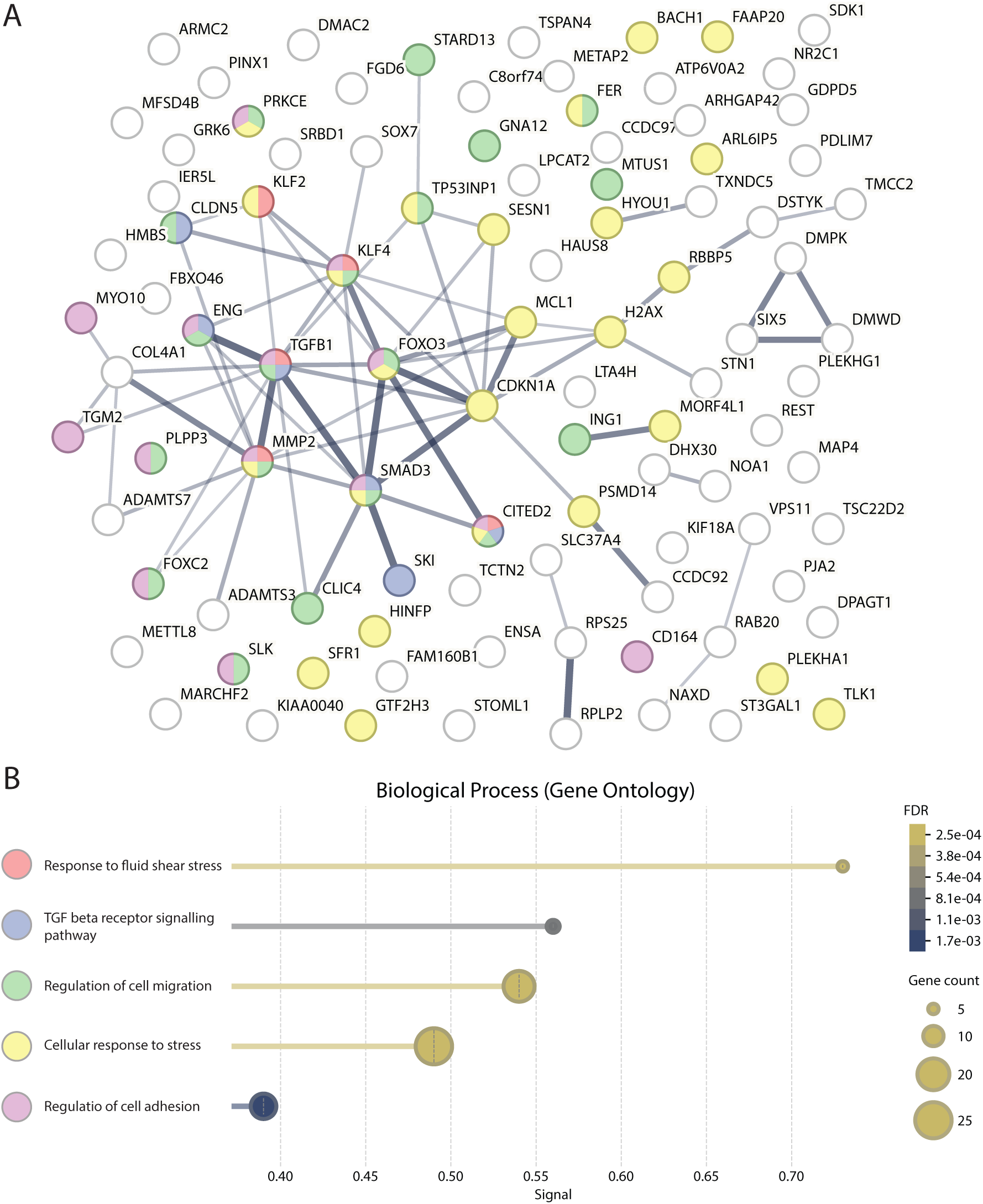
Analysis of the endothelial enhancer-promoter map for CAD genetics. A. String diagram showing genes linked by MCC to a HUVEC enhancer containing a candidate causal CAD SNP. Interactions between a captured MCC enhancer and gene promoters were called using the bespoke MCC peakcaller MCC_LoTron. Gene promoters were defined using REgulamentary. Nodes which are enriched for specified GO terms are highlighted by colour. The thickness of the connecting line corresponds to the strength of the interaction evidence between two nodes. B. Gene Ontology analysis (biological processes enrichment) of the MCC derived gene network.

### Allelic skew shows causality of SNPs

Having linked CAD associated enhancer SNPs to output genes using MCC, we next investigated specific loci where it is possible to demonstrate causality. SNPs which alter enhancer function are thought to change transcription factor binding occupancy. This can be directly detected in MCC datasets by examining the MNase cutting pattern around the SNP. Where the donor cells are heterozygous for the candidate causal SNP, the MNase cutting pattern can be directly compared between the two alleles within the same cell. For example, at the *MMP2* locus HUVEC donor 2 is heterozygous for the candidate causal SNP rs80345658 (Figure 5A). The MCC signal was split into two separate tracks, revealing a significantly stronger interaction with the promoter of *MMP2* from the rs80345658 enhancer containing the A allele compared to the G allele (Figure 5B). This observation indicates that the A allele strengthens the enhancer leading to a more active *MMP2* promoter. Allele specific analysis of the enhancer shows a markedly different footprinting profile around the rs80345658 SNP (Figure 5C). This is consistent with preferential binding of a transcription factor to the A allele compared to the G allele, demonstrating causality. Analysis of the sequence around the SNP indicates that the G allele may disrupt a SMAD1/4 binding motif (Figure 5D). All allele specific data for heterozygous SNPs can be viewed here: https://genome-euro.ucsc.edu/s/mdehxmb7/CAD_HUVEC_MCC_allelic

**Figure 5.**
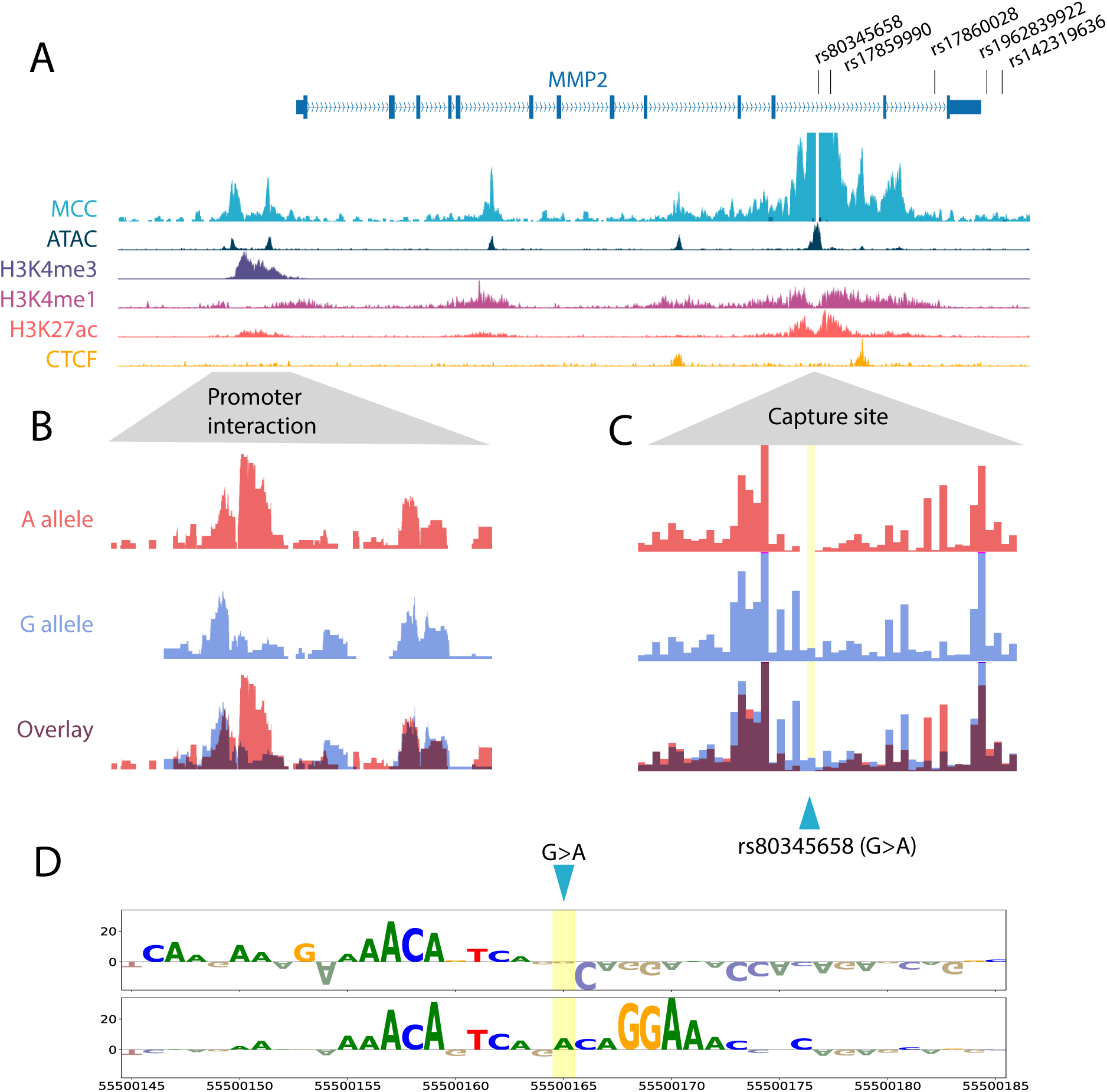
Allelic MCC shows causality of enhancer SNPs. (A) MCC capture probes were designed across candidate causal SNP rs80345658 which is found in an enhancer in endothelial cells. MCC revealed an enhancer-promoter interaction from this SNP to teh MPP2 gene promoter. The MCC tracks were split by whether the signal was derived from the allele containing an “A” or a “G” at the SNP position. By doing so it was possible to compare (B) the promoter interaction from the enhancer and also (C) the enhancer footprint around the SNP site. (D) REnformer prediction of candidate causal SNP rs80345658 demonstrates that the A-allele creates a SMAD binding motif.

### POINTseq measures allele specific gene output

To determine whether the captured enhancer SNPs lead to altered gene output we combined long read genome sequencing with strand specific POINTseq. Long read sequencing was used to phase SNPs and reconstruct alleles for each of the HUVEC donors. We then used POINTseq to measure allele specific transcription of genes linked to candidate causal enhancer SNPs by MCC (Figure 6A). POINTseq affords an advantage over traditional RNAseq (polyA+), and even polyA-RNAseq, in terms of intron coverage (Figure 6B). Phasing SNPs between enhancers and promoters enabled the direction of change to be linked between output genes and CAD enhancer SNPs. To avoid alignment bias to the reference genome, all reads were aligned to reconstructed alleles 1 and 2 (for the respective HUVEC donors), as well as the GRCh38 reference genome. The allelic skew was then measured for each donor specific heterozygous SNP within the transcriptional body of the MCC linked output gene. At each SNP position, the alignment with the least skew was taken, before averaging across the gene. This conservative approach is likely to underestimate the true magnitude of the transcriptional skew at each candidate effector gene, however the confidence with which we can claim statistical significance is strengthened. In total, significant transcriptional skew was detected in 18 MCC linked genes (Figure 6C). Notably, a number of these genes are involved in TGF-β mediated responses including *SMAD3*, *MMP2*, *SKI*, and *FOXO3*. The detected skew for all genes linked by MCC to a heterozygous enhancer SNP in HUVEC donors 1-3 is summarised in Supplementary Table 4.

**Figure 6.**
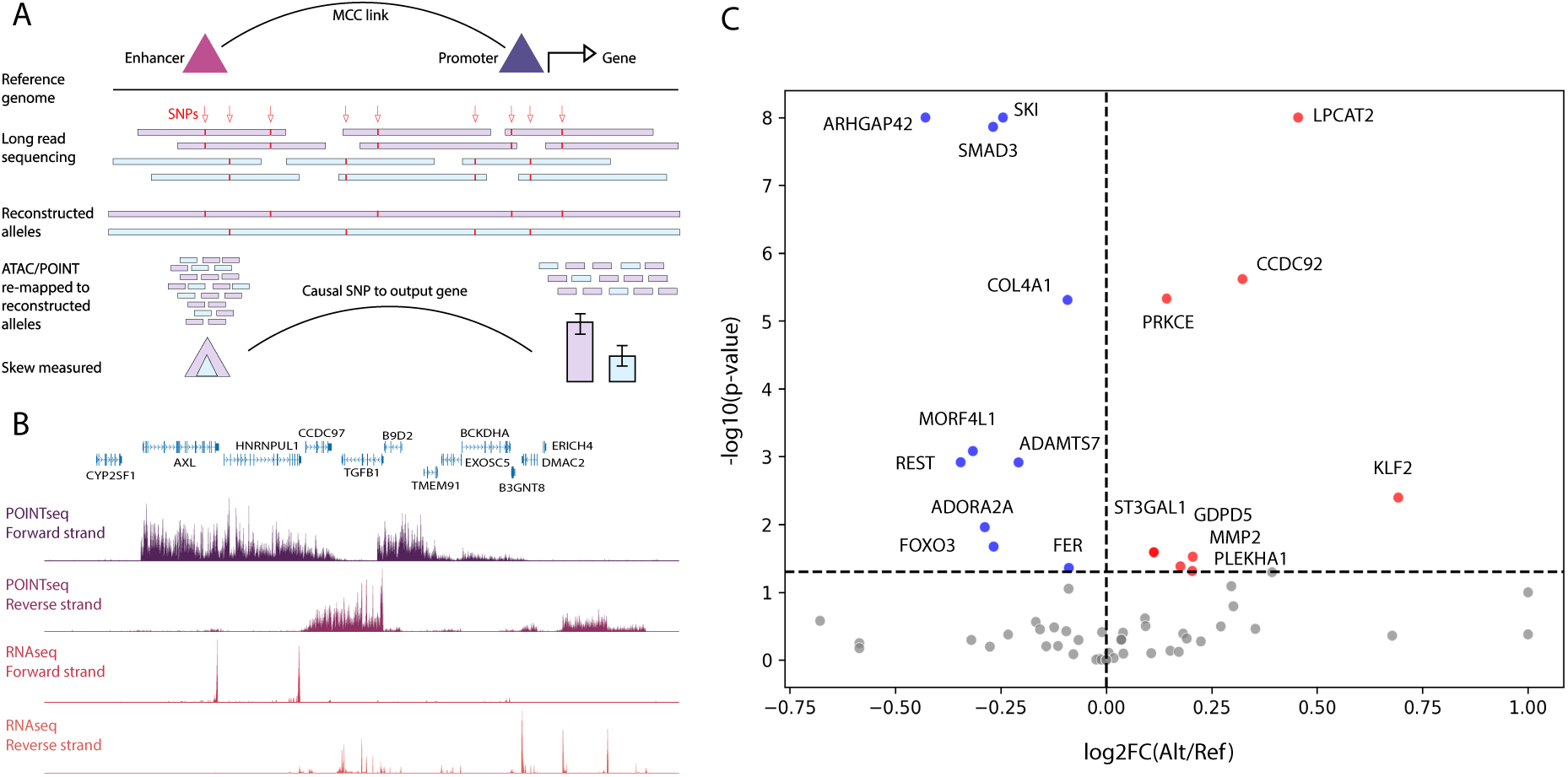
POINTseq detects the changes in MCC linked output genes. A. Schematic overview of experimental approach. MCC links candidate causal enhancer SNPs to the promoters of output genes. Long read sequencing is used to phase SNPs and reconstruct the alleles from the donor cells. ATACseq and POINTseq data is re-mapped to the reconstructed alleles and reads are assigned to the allele they are derived from based on corresponding SNPs. The amount of signal coming from each allele can then be measured and compared, demonstrating the effect of a casual SNP in an enhancer on gene transcriptional output. B. Example POINTseq data tracks compared to traditional RNAseq. POINTseq, polyA-, and polyA+ RNAseq were performed on 3 HUVEC donors. Reads are split between forward and reverse DNA strand, mapped to the hg38 reference genome, and displayed as bigwig files. POINTseq offers far greater transcriptional gene coverage as introns are retained. D. Plot of POINTseq measured allelic skew in genes linked by MCC to candidate causal enhancer SNPs. Direction of change is displayed with respect to the reference allele versus the alternate allele (x-axis). Significance (log10 p-value) is displayed on the y-axis. Detected significant genes are labelled.

## Discussion

The primary aim of this study was to develop a computational and experimental pipeline to resolve GWAS associations, identifying key effector genes and pathways for therapeutic development. We chose to develop these approaches using CAD genetics because of the mechanistically complex nature and major global impact of this disease. To do this we combined ML and genomic technologies which we believe constitutes a blueprint for future endeavors to elucidate mechanisms underlying genetic associations with disease traits and expedite the discovery of new therapeutic targets. We began by curating a list of lead SNPs for CAD associations, before imputing all potential causal SNPs in strong linkage disequilibrium with these lead SNPs. We trained the ML platform REnformer with publicly available scATAC data to prioritise both candidate causal SNPs and coronary artery cell types. We selected endothelial cells as a highly important and tractable cell type for validation and mechanistic exploration. We performed MCC to link these candidate causal SNPs to output genes and took advantage of heterozygous SNPs in the HUVEC donors to look for allelic skew to infer changes in transcription factor binding and enhancer-promoter interaction. Lastly, we employed long-read sequencing and POINTseq to link candidate causal SNPs to altered transcriptional output of key effector genes.

We believe these technologies offer significant advantages over other existing functional genomics approaches. The ML tool REnformer allows for the *in silico* testing of all candidate variants across all potential effector cell types simultaneously. The ability to simultaneously resolve causal variant and effector cell type massively reduces the experimental complexity required to find target genes offering huge advantages over traditional fine-mapping variant prioritisation tools.

The combination of allele-specific MCC with genome-phased POINTseq allows for direct observation of SNP causality. Causal variants in cis-regulatory elements such as enhancers function by altering transcription factor binding dynamics and these changes can be observed in the MCC footprints. MCC also provides the highest resolution enhancer-promoter interaction maps, providing significant advantages over Hi-C, Capture-C, and Activity-by-Contact (ABC) models when linking causal enhancer SNPs to target genes. Furthermore, allele-specific changes in enhancer-promoter interactions can be directly observed, linking altered transcription factor binding to altered enhancer function. Because the MCC assay is conducted on native chromatin, there is a significant detection advantage over massively-parallel-reporter-assays (MPRAs) which lack 3-dimensional genomic context, and CRISPRa/i based perturbation studies which do not recapitulate the subtle effects of single base pair changes.

We intend the datasets generated in this study to be used as a resource for other researchers to exploit to determine new targets for the treatment of CAD. Researchers with an interest in endothelial biology can search for causal SNPs and output genes in their loci of interest by browsing the UCSC session (https://genome-euro.ucsc.edu/s/mdehxmb7/CAD_HUVEC_extended_data). Here, researchers can evaluate the evidence for a candidate causal SNP themselves to pick targets and pathways for validation in functional assays and model systems.

The genes and pathways identified in this analysis build on the considerable efforts by other groups to resolve cardiovascular genetics. For example, we identify the transcription factors *KLF2* and *KLF4* which are master regulators of endothelial gene programs in the CCM pathway recently highlighted in Perturb-seq studies of teloHAECs ^23^. That study also identified that *CCM2* inhibits *RHOA* to modify actin remodeling, and our study identifies the *RHOA* inhibitor *ARHGAP42*. Increased expression of the lipid phosphate phosphatase gene *PLPP3* has been shown to be atheroprotective in mouse models and human aortic endothelial cells exposed to disturbed flow in vitro ^24,25^. We identified two candidate causal SNPs in this locus located in endothelial enhancers. Through MCC experiments we were able to show that the rs11206803 enhancer, but not the rs56348932 enhancer, interacts with the promoter of *PLPP3*. This is an example of how the data in this study can be used to resolve the genetic mechanism underlying a known CAD modifier gene. We also identified the lead SNP rs582384 in an endothelial enhancer interacting with the promoter of *PRKCE*. This kinase, which is known to be involved in the development of cardiac hypertrophy, may also play a role in the progression of CAD and is a potentially tractable drug target (TargetDB).

While the identification of individual target genes is an important step forward, doing this at scale is critical to gain a picture of the underlying biology altered by the genetics. Due to the number of associated genes identified in our study we were able to demonstrate the prominence of endothelial shear stress pathways in CAD genetics. While other processes including oxLDL signalling and nitric oxide production have long been known to affect CAD progression, recent studies have also highlighted the importance of mechanosensing and fluid shear stress using in vitro model systems. Fluid shear stress induces endothelial *TGFB1* production ^26^, and we identified an enhancer containing a CAD SNP that interacts with the promoter of *TGFB1*. The role of *TGFB1* in CAD is multifaceted with both atheroprotective and atherogenic effects. *TGFB1* can activate *MMP2* through SMAD-dependent pathways. In this study we identified the SNP rs80345658 which increases the activity of an intronic enhancer that interacts with the promoter of *MMP2*. In this biological context an increase in *MMP2* expression would be consistent with promotion of the initial development of atherosclerotic plaques through degrading extracellular matrix in vessel walls, as well as plaque instability through degradation of the fibrous cap.

Our aim was to generate a combined platform of highly scalable approaches to accelerate translating the type of precise genes and pathways identified in this study into therapeutic strategies using relevant disease models. The experimental approach described in this manuscript should also be extended to other tractable CAD cell types such as pericytes and smooth muscle cells. The genes and pathways from multiple cell types can then be integrated to identify novel therapeutic targets and reduce the global impact of the primary cause of death worldwide. This approach could also be replicated to discover causal SNPs, output genes, and tractable pathways in other diseases with GWAS data.

## Supplementary data

MCC probe design: https://mlv.molbiol.ox.ac.uk/projects/multi_locus_view/8712

MCC viewpoints for 94 enhancers: https://genome-euro.ucsc.edu/s/mdehxmb7/CAD_HUVEC_extended_data

Allele-specific MCC tracks: https://genome-euro.ucsc.edu/s/mdehxmb7/CAD_HUVEC_MCC_allelic

Supplementary Table 1:

SNP curation from Aragam et al. The index_rsid are either genome-wide significant, 1% FDR or FGWAS from endothelial cells or smooth muscle cells. Proxies for the index SNP were collated using LDLinkR with r2>0.8.

Supplementary Table 2:

REnformer ML predictions for curated candidate CAD SNPs across 9 primary coronary artery cell-types (endothelial, fibroblast, macrophage, mast, pericyte, B cell, smooth muscle 1&2, T cell) and HUVECs. Predictions can be filtered by Lead SNP (“Region_index”).

Supplementary Table 3:

MCC enhancer-promoter map for HUVEC donors 1-3.

Supplementary Table 4:

POINT-seq derived allelic skew of MCC-linked candidate target genes.

Supplementary Table 5:

Full gene ontology analysis of MCC-linked candidate target genes.

## Methods

### Variant selection

FGWAS ^27^ software was used to identify functional variants associated with CAD ^2^ using smooth muscle cell (GSE72696) & endothelial cells (GSE126196) chromatin data. Summary statistics were downloaded from CARDIoGRAMplusC4D website and integrated with chromatin data to perform genome-wide FGWAS analysis. Variants with PPA>5% were selected along with all genome-wide significant variants and 1% FDR independent SNP list. 1000G reference was used identify proxies (r2>0.8) for the 1289 selected variants to use all variants in the haplotype block.

### Cell culture

Single donor human umbilical vein endothelial cell (HUVEC) lines were purchased from Promocell (C-12200) or were isolated from umbilical cord tissue samples provided by the Anthony Nolan Trust, who received ethical approval from the East Midlands-Derby, UK Research Ethics Committee and written, informed consent from the donor’s parents. HUVECs were isolated from the umbilical vein as described previously ^28^. Briefly, the vein was cannulated and one end clamped. The vein was flushed with PBS followed by air. 0.5mg/ml collagenase solution was added to the vein and the open end was clamped. After massaging the tissue sample, the tissue was incubated at 37°C for 15 minutes. The collagenase solution (containing the HUVECs) was collected in a centrifuge tube and the vein washed with PBS to collected any remaining cells, which was also added to the centrifuge tube. The cell suspension was centrifuged at 500xg for 5 minutes and the cell pellet was resuspended in complete HUVEC media and transferred to a T75 flask coated with 0.2% gelatine. The cells were cultured in complete HUVEC growth media (M199 supplemented with 15% foetal calf serum [FCS], 1% penicillin/streptomycin, 10U/ml heparin, 4.5µg/ml endothelial cell growth supplement [ECGS], 5ng/ml human FGF-acidic and 2.5µg/ml thymidine) at 37°C/5% CO_2_. Cells were harvested for experiments between passages 5-7.

### Machine Learning

#### Training data

The raw scATAC-seq sequencing data of coronary artery (CAD) samples from Turner et al ^4^ were aligned to the hg38 reference genome using bwamem2 (v2.2.1). Samples from different patients were pre-processed separately. Low-quality reads were filtered out, duplicates were removed using SAMtools (v1.17). scATAC-seq reads from different samples were merged and then quality control filtering, dimensionality reduction and clustering was performed using ArchR (v1.0.2). Before quality control filtering, the total number of cells was 130,892. For quality control filtering, cells with a TSS enrichment score of ≥7 and a unique barcode count of ≥10,000 were retained, resulting in the final number of cells being 21,275. The annotation method and UMAP embeddings from Turner et al ^4^ were used to assign cell type identities to the 21,275 cells and visualize them on the same UMAP. Pseudo-bulk ATAC tracks for each cell type were reconstructed from the scATAC reads from each of the retained cells within each cluster. These Pseudo-bulk ATAC tracks were used for fine-tuning REnformer.

#### Evaluation

To evaluate predictive performance across heterogeneous cell types and across genomic regions with widely varying signal intensity, we adopted the three normalised root mean square error (NRMSE) metrics introduced in the REnformer benchmark (Ref REnformer). All metrics derive from the base RMSE computed per region i and cell type (track) j. For a genomic region containing n=896 bins, the RMSE is defined as:

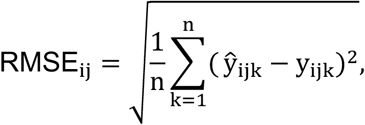

where 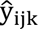 and y_ijk_denote the predicted and observed signal, respectively.

Because different cell types exhibit distinct dynamic ranges, the RMSE was normalised in three complementary ways following the methodology described by Riva et al ^19^:

1. Variance-normalised RMSE (NRMSE₁; “Var”)

This metric scales RMSE by the standard deviation of the ground-truth signal within the region:

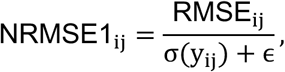

where σ(y_ij_)is the standard deviation across the n bins, and ɛ = 10^−8^prevents division by zero for constant-valued regions.

2. Min–max-normalised RMSE (NRMSE₂; “Inter”)

To capture error relative to the full amplitude of the observed signal, RMSE is normalised by the range:

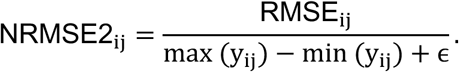

This metric is sensitive to dynamic range and is useful for tracks with pronounced peaks.

3. Inter-quartile-range-normalised RMSE (NRMSE₃; “Diff”)

To provide a robust normalisation that reduces sensitivity to extreme values, RMSE is divided by the interquartile range:

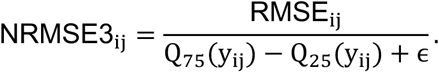

This is particularly informative for highly skewed or sparse signals.

Regions in which the target signal consisted entirely of zeros were excluded, following the REnformer procedure, to avoid undefined variance-based denominators. Remaining regions were evaluated independently for each metric, and distributions were summarised across cell types using the log-transformed NRMSE values.

Fine-Tuning REnformer for Open Chromatin and Variant Prediction.

We adapted the Enformer architecture to train REnformer ^19^, a sequence-based deep learning model specialised for predicting open chromatin profiles and regulatory variant effects. REnformer retained Enformer’s hybrid convolution-transformer backbone, which combines local motif detection with long-range attention to capture distal regulatory dependencies across ∼200 kb genomic contexts. The REnformer model was initialized from pre-trained Enformer weights and subsequently fine-tuned on specific single cell-type dataset, which focuses on chromatin accessibility signals. This fine-tuning enabled REnformer to recalibrate sequence representations toward cell-type specific regulatory architectures while maintaining the general regulatory grammar learned from the multi-cell-type Enformer model. Variant effect predictions were obtained by comparing REnformer’s output for reference and alternate alleles, providing base-resolved estimates of how sequence variants modulate chromatin accessibility.

#### Feature attribution

Model interpretability was assessed using gradient × input attribution which quantifies the contribution of each nucleotide to the predicted chromatin accessibility signal ^29,30^. This method provides base-resolution insight into the sequence features driving model predictions and highlights regions with strong positive or negative influence on accessibility. The resulting attributions frequently coincide with known transcription-factor binding motifs indicating that REnformer captures biologically meaningful regulatory patterns.

We did not reanalyse transformer attention maps, as prior work from the Enformer model demonstrated that attention patterns reliably capture distal enhancer-promoter relationships. Given this established finding, and because gradient × input offers a more direct and fine-grained measure of sequence-level feature importance with lower computational overhead, we focused our interpretability analyses solely on gradient-based attributions. Collectively, these results confirm that REnformer effectively learns both local motif structure and long-range regulatory dependencies underlying open chromatin across multiple cell types and with or without variant effect predictions.

### Binding motif analysis

Transcription factor binding motifs were identified using Tomtom in MEME Suite version 5.5.9 ^31^. The query sequence was 41 basepairs around the variant (+/- 20 from variant location) and the search database was HOCOMOCO Human v11.

### ATAC sequencing

ATAC sequencing protocol was adapted from the published protocol ^27^ with minor modifications. 200,000 cells were harvested per HUVEC donor and split across 3 technical replicates (approximately 66,000 cells per replicate) into ice cold lysis buffer (10 mM Tris-HCL, 10 mM NaCl, 3 mM MgCl_2_, 0.1% Igepal CA-630). Nuclear pellets were washed with ice cold PBS to remove mitochondrial DNA. Tagmentation was performed with Tn5 transposase from Illumina (15027865) and libraries were sequenced on a NextSeq 550. Data was processed and analysed using CATCH-UP ^32^.

### Regulatory genome annotation

Genomic cis-regulatory elements were annotated using REgulamentary ^33^. In-house ATAC sequencing data was used in combination with publicly available ChIP-seq for H3K4me1, H3K4me3, H3K27ac, and CTCF, downloaded from ENCODE. All data was preprocessed using CATCH-UP before analysis with REgulamentary.

### Micro Capture-C

Micro Capture-C experiments were performed according to the published protocol ^21^. Briefly, HUVECs were fixed with formaldehyde before permeabilization with digitonin and digestion with MNase. Chromatin was re-ligated before DNA purification and sonication to around 200bp. 6 independent MNase digests were performed for each HUVEC donor and indexed separately before pooling for double capture with 120bp biotinylated DNA probes. Captured libraries were PCR amplified and sequenced on a NovaSeq 6000 with 1 million reads per digest per capture probe. Raw sequencing data was processed according to the published protocol (https://github.com/jojdavies/Micro-Capture-C) to generate alignment files for each digest; all 6 digests for each HUVEC donor were then merged to create genome coverage tracks for each capture probe (i.e. viewpoint). Genomic interactions with the capture point were then called using MCC_LoTron, a bespoke peak caller developed for MCC data. Allele-specific MCC was performed and analysed as previously described ^34^.

### Long read genome sequencing

Genomic DNA was extracted using Nanobid CBB kit (102-572-200) and sequenced on a PacBio Sequel IIe. Raw, single-end reads in unaligned bam format were first aligned with PBMM2 v1.13.1 using the CCS preset and the sort option, using 32GB of memory and 30 threads. These were aligned to a pbmm2 index created using the UCSC hg38 genome fasta, in the minimap2 standard MMI format. The resulting aligned bam file was indexed with samtools v1.20 using 30 threads immediately thereafter. SNPs were called using Deepvariant with NVIDIA Clara Parabricks version 4.3.0-1 docker image, using 14 threads per stream, 2 streams per GPU, 2 GPUs (A5000 ampere generation), 100000 maximum reads per partition, 20000 partition size, using the PacBio mode, and the prealign helper thread option. The Deepvariant VCF was then annotated using bcftools into BCF format, then converted to VCF with the bcftools convert utility. The VCF was then compressed with bgzip using 32 threads and tabix indexed (both using htslib v1.17). Whatshap phasing occurred using the indels, include homozygous, ignore read groups, and distrust genotypes options against the hg38 genome. The phased VCF was then again compressed with bgzip using 32 threads and tabix indexed. Using this phased VCF, the aligned bam was haplotagged using the option to tag supplementary reads and to ignore read groups. The resulting haplotagged bam was sorted by the HP tag using 56 threads and 4GB memory per thread. Whatshap version 2.4.dev6+g96e2911 (installed from GitHub at the time of the g96e2911 commit), samtools v1.21, and bcftools v1.20 were used for the preceding step. The pipeline was run locally, using snakemake 8.15.2.

### POINT-seq

Point-seq was performed as previously described with some modifications ^22^. Cells were lysed and nuclei extracted as previously described before resuspension of the chromatin pellet in ice cold NUN-1 buffer. Resuspended chromatin was then treated with NUN-2 buffer containing 3% Empigen and incubated on ice for 10 minutes to precipitate the chromatin. Samples were centrifuged at 400 xG for 1 minute at 4°C to pellet the chromatin which was then washed twice with PBS. Pelleted chromatin was then resuspended in 200 ul of DNase Turbo buffer with 3 uL of DNase Turbo enzyme (AM2238) and incubated for 15 minutes at 37°C in a thermoshaker at 1000 rpm. 5 uL of proteinase K (P8107S) was added to each sample before a further 15 minutes incubation at 37°C in a thermoshaker at 1000 rpm, followed by phenol-chloroform extraction. Purified nucleic acids were then washed with 70% cold ethanol before a second round of DNase Turbo digestion for 15 minutes and POINT RNA was finally isolated using Trizol and chloroform. Purified POINT RNA was prepared for sequencing using NEBNext Ultra II directional RNA Library Prep Kit (E7760) with depletion of ribosomal RNA and sequenced on a NextSeq 550 (150 bp paired-end).

### Detection of allelic skew

We developed a bespoke analysis pipeline for the detection of allelic skew in POINT-seq data. The code this pipeline is available here: https://github.com/Genome-Function-Initiative-Oxford/MCCGeneSkew. For each donor the POINT-seq fastq files were aligned using CATCH-UP pipeline to the standard hg38 genome, as well as to the personalised genomes of each derived from the Pac-Bio long read sequencing data (bcftools). The resulting BAM files were split into forward and reverse strand to remove interference from overlapping transcription on opposite strands. Pac-Bio phasing of individual donors was used to determine which heterozygous SNPs were in phase with MCC-captured enhancer SNPs. At each heterozygous SNP position within an MCC defined target gene read counts were generated for each allele using pysam pileup.

Allele-specific read counts were obtained from the bam files using pyliftover. Statistically significant differences between the read counts on the reference and alternative allele were detected using the published WASP ^35^ beta-binomial model using ‘combined_test.py’. Only the allelic skew (--as-only) component is used due to other requirements being satisfied due to PacBio phasing with read counts obtained on personalised genomes.

## Data Availability

Raw data is available here: GSE304307

## Acknowledgements

This work was funded by grants from the BHF-DZHK (SP/19/2/344612), Wellcome Trust (225220/Z/22/Z), and Medical Research Council (MC_UU_00016/14, MC_UU_00029/3). A.G. has received support from the BHF, European Commission [LSHM-CT-2007-037273, HEALTH-F2-2013-601456], BHF-DZHK (SP/19/2/344612). H.W. is supported by CureHeart, the British Heart Foundation’s Big Beat Challenge award (BBC/F/21/220106). D.G.M, S.Y and T.R.W received funding from the British Heart Foundation (RG/16/13/32609, RG/19/9/34655, PG/16/9/31995, PG/18/73/34059, and SP/19/2/344612). D.G.M is supported by BHF Research Excellence Award (RE/24/130031) and (AA/18/3/34220).

We thank Fatin Al-Janabi for her contribution to the HUVEC collection.

---

**Supplementary Figure S1.**
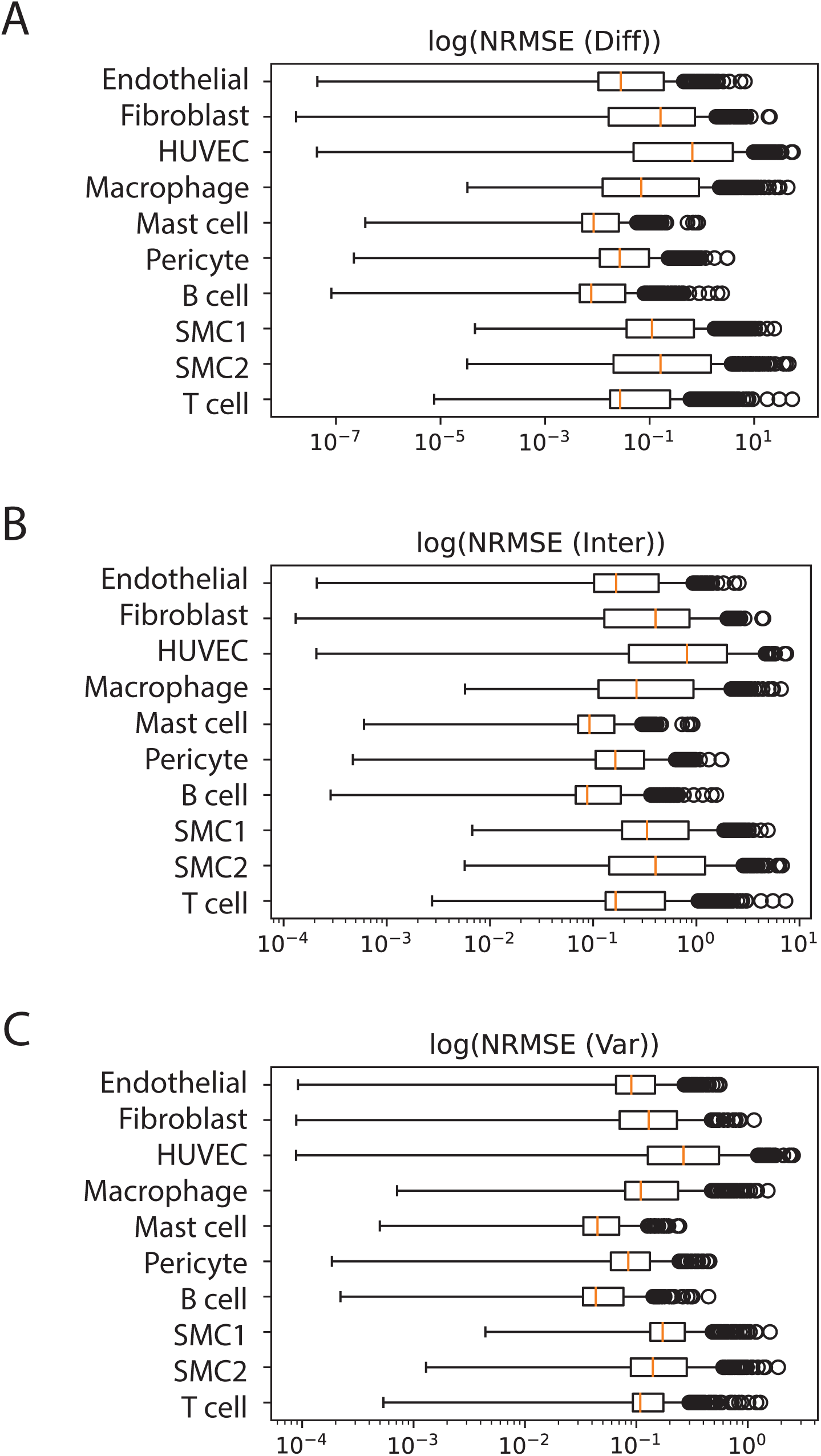
Cell-type specific distributions of log-normalised RMSE metrics across REnformer predictions. Boxplots show the distribution of log(NRMSE) metrics across the hold out test genomic regions for each of 10 representative cell types, evaluated using the three normalised RMSE metrics introduced in the REnformer benchmarking framework. From left to right, panels summarise: (A) NRMSE(Var): RMSE normalised by the per-track standard deviation; (B) NRMSE(Inter): RMSE normalised by min–max range; and (C) NRMSE(Diff): RMSE normalised by the inter-quartile range (IQR). Each point represents one genomic region, and boxplots illustrate median, interquartile ranges, and extreme values on a log scale. Together, these metrics quantify error magnitudes under different normalisation regimes, allowing consistent comparison of prediction difficulty across both cell types and dynamic range of signal. Across all three normalisation schemes, the consistently low median log(NRMSE) values and compact interquartile ranges indicate robust and accurate predictive performance, demonstrating that the model generalises well across diverse cell types and genomic contexts.

**Supplementary Figure S2.**
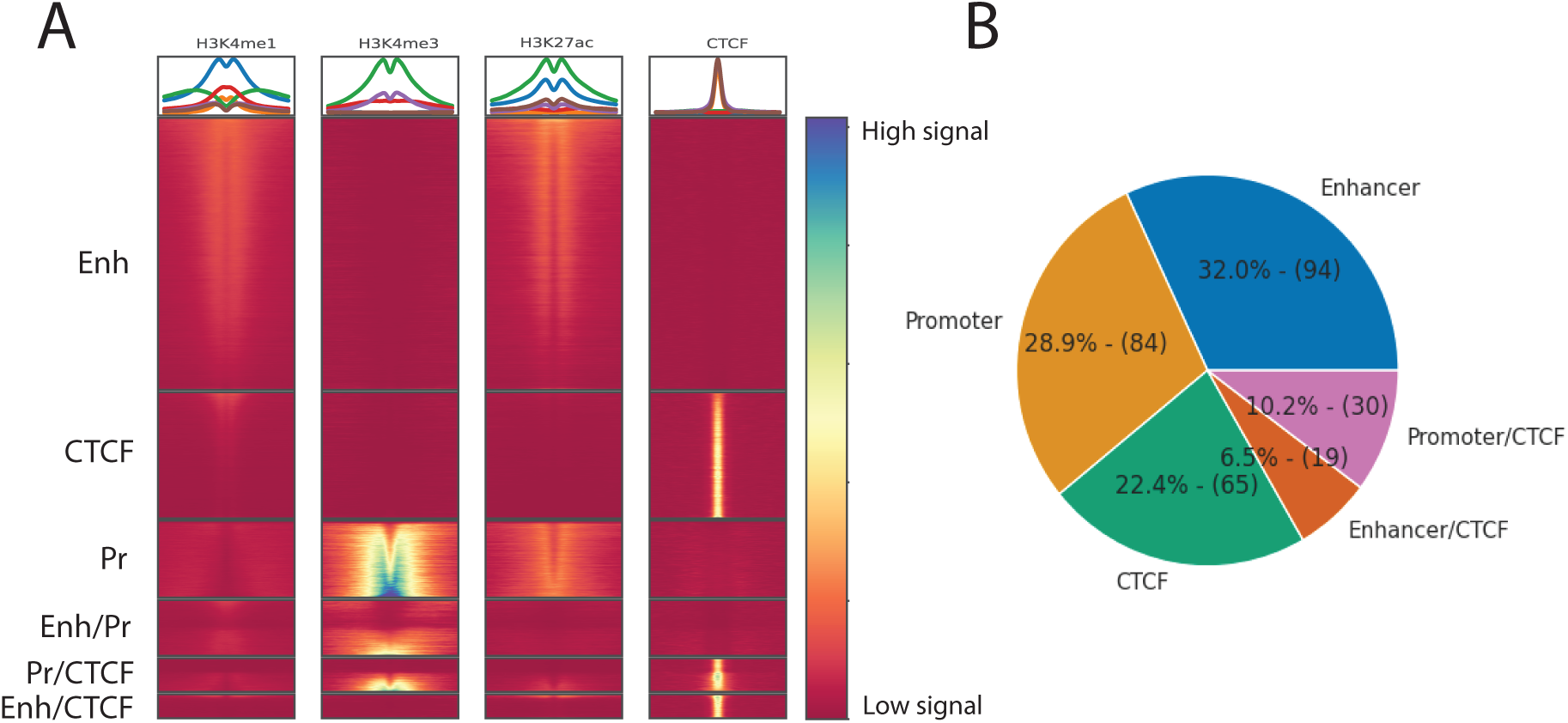
A. Genome regulatory elements including enhancers (Enh), CTCF sites, and promoters (Pr) were determined for HUVECs using REgulamentary. B. Chart showing the number of intersections between REgulamentary defined regulatory elements and candidate causal variants.

